# Exploring Data Augmentation for Stroke Severity Prediction: How Generative AI Enhances EEG Signal Classification

**DOI:** 10.64898/2026.11.09.26367632

**Authors:** Md. Sadman Sakib Sarkar, Md. Tanvir Hossain Saon, Sarnali Basak

## Abstract

In clinical datasets, data augmentation is often avoided because it can remove important unusual values, known as outliers, which are important for understanding diseases accurately. However, clinical EEG datasets often suffer from two key limitations: small sample sizes and class imbalance, which significantly reduce classification performance. To over-come these challenges, this study applies generative AI-based data augmentation techniques with a carefully designed hybrid pipeline that effectively enhances performance even on limited EEG data. Using EEG signals from 50 brain stroke patients [1], we investigate both traditional oversampling methods (SMOTE and ADASYN [2]) and advanced generative models, including WGAN and WGAN-GP. Our proposed pipeline involves pre-processing EEG recordings, extracting comprehensive statistical and frequency-domain features, applying feature reduction, and evaluating multiple machine learning classifiers. Experimental results show that the combination of SelectKBest feature selection, SVM classifier, and WGAN-GP augmentation achieves the highest classification accuracy of 72%, outperforming other methods. This work introduces a novel hybrid approach for EEG-based stroke severity classification, demonstrating that generative augmentation, when integrated properly, can significantly improve performance on small clinical datasets. To the best of our knowledge, this combination of techniques has not been explored before on any dataset, providing a solid foundation for future research in automated EEG-based stroke diagnosis.

## I. Introduction

Stroke remains one of the leading causes of long-term disability and death worldwide, posing a significant burden to healthcare systems [3]. Accurate classification of stroke severity is crucial for timely treatment decisions and improved recovery outcomes. Electroencephalography (EEG) is a widely used tool for assessing brain activity and diagnosing neurological conditions, including stroke. However, EEG-based automated stroke severity classification faces two primary challenges: limited data availability and severe class imbalance, which can negatively impact machine learning model performance. Imbalanced datasets, where certain classes (such as mild or severe) are underrepresented, often lead to biased predictions and reduced model reliability. While data augmentation is a common strategy to address class imbalance, in clinical datasets it is typically approached cautiously because it may eliminate critical outliers or distort rare but diagnostically significant patterns, particularly in disease analysis where such extremes are vital. Nonetheless, in scenarios like stroke EEG analysis, where patient samples are limited, data augmentation becomes a practical necessity to enhance classification performance.

To tackle these challenges, this study proposes a novel hybrid pipeline integrating feature selection with generative augmentation to improve stroke severity classification. Recent research shows that Generative Adversarial Networks (GANs) can generate realistic synthetic EEG signals [5], with advanced variants like WGAN-GP achieving superior performance in tasks such as seizure detection [6]. Inspired by these advancements, this work applies four augmentation techniques—SMOTE, ADASYN, WGAN, and WGAN-GP—on EEG data from 50 stroke patients [1], combining preprocessing, statistical and frequency feature extraction, dimensionality reduction via SelectKBest, and SVM classification. Experiments demonstrate that this hybrid approach, especially the combination of WGAN-GP, SelectKBest, and SVM, achieves a classification accuracy of 0.72, outperforming traditional oversampling methods and baseline models. This research highlights the potential of generative AI to address class imbalance and improve classification accuracy in EEG-based stroke severity assessment, offering a framework to aid stroke diagnosis and treatment.

## II. Literature Review

Generative Adversarial Networks (GANs) have shown great promise in addressing EEG data scarcity and class imbalance, improving classification outcomes in various domains.Hortal and Bhat [7] used WGAN-GP on the DEAP dataset for emotion recognition, achieving classification gains of up to 17.5% through GAN-based data augmentation. Similarly, a TGAN model enhanced seizure detection using thalamic SEEG data, improving accuracy and sensitivity significantly [8].Conditional WGAN was explored in [9] to augment EEG data, enhancing DNN classification by 6.5% (arousal) and 3.0% (valence). ESC-GAN [10] further improved performance in class-imbalanced emotion recognition tasks by generating diverse and boundary-aware samples across DEAP, AMIGOS, and SEED datasets.

Augmentation has also aided clinical classification tasks. Ullah et al. [11] combined a pyramidal CNN with augmentation techniques, achieving 99.1% accuracy in epilepsy detection. LS-GAN improved small medical dataset classification in [12], especially for SVM. Differential privacy in synthetic EEG data generation was explored in PATE-GAN [13], balancing data utility and confidentiality. A broader survey of augmentation methods [14] highlighted GANs’ strengths in small, imbalanced datasets, particularly in medical imaging.Hartmann et al. [15] introduced EEG-GAN using WGAN-GP to synthesize realistic EEG signals for BCI applications. Studies on motor imagery (MI) classification [16], [17] emphasized CNN-based architectures’ efficiency, with data augmentation boosting real-time neuroprosthetic control.WGAN-GP’s superiority in seizure detection was again shown by Abou-Abbas et al. [6], with accuracy rising from 86% to 91.73% after augmentation. Similarly, L-C-WGAN-GP [18] and DCGAN-GP [20] models significantly improved MI EEG classification by generating high-fidelity synthetic signals.Several studies [19], [21] reinforce the effectiveness of deep learning for EEG signal classification, and highlight augmentation’s role in increasing BCI accessibility. Finally, GANs have also been effective in denoising EEG signals for real-world applications [22], enhancing signal quality and system robustness. These studies collectively demonstrate the growing impact of generative models in EEG research, particularly in handling limited, imbalanced data—central to our work on brain stroke severity classification.

## III. Methodology

This section outlines the methodology for EEG-based stroke severity classification, integrating feature engineering, dimensionality reduction, classification, and data augmentation to address class imbalance. The workflow, depicted in Figure 1, encompasses raw data preprocessing, feature extraction, reduction, model training, and evaluation within a 5-fold cross-validation framework.

**Fig. 1.**
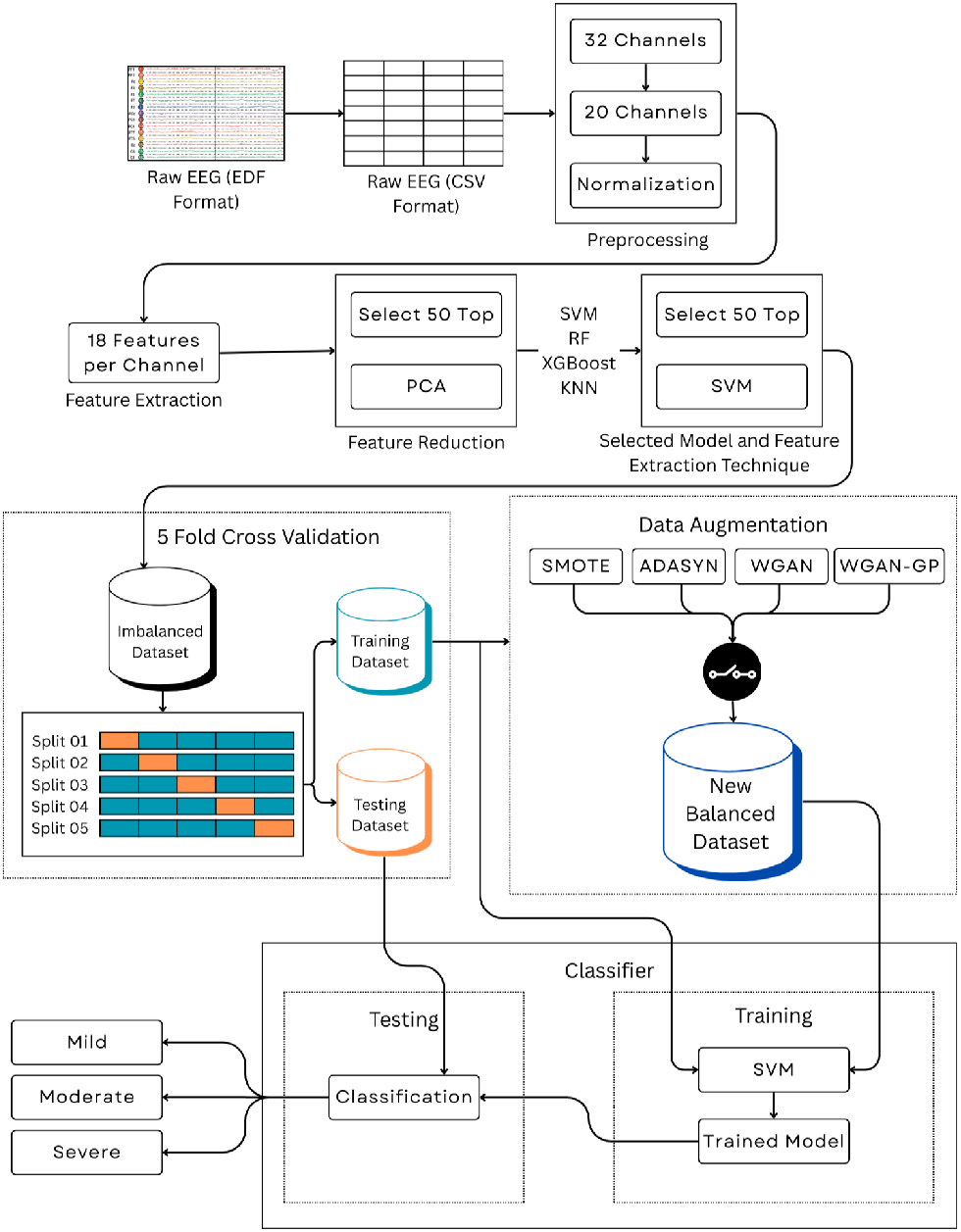
Flowchart of the proposed EEG-based stroke severity classification pipeline.

### A. Dataset and Preprocessing

The study utilizes an open-access EEG dataset from Liu et al. [1], comprising signals from 50 acute ischemic stroke patients, recorded in European Data Format (EDF) with approximately 160,000 samples per subject across 32 channels based on the 10–20 international standard. The National Institutes of Health Stroke Scale (NIHSS) scores categorize stroke severity into mild,moderate, and severe.

Preprocessing involved converting EDF files to CSV format for compatibility with Python-based tools. A variance-based channel selection retained the top 20 channels with the highest average variance across all subjects, calculated as:

,
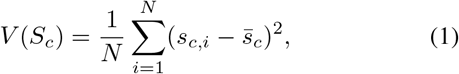

,
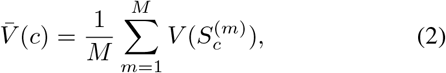

where *S*_*c*_ represents the signal for channel *c, s*_*c,i*_ is the *i*-th sample, 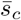 is the channel mean, *N* = 160, 000 is the number of samples per file, and *M* = 50 is the number of subjects. Channels with standard deviation below 10^*™*6^ were excluded to eliminate low-variance signals.

The selected channels were normalized using the Standard-Scaler from scikit-learn, transforming signals to zero mean and unit variance:

,
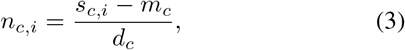

,
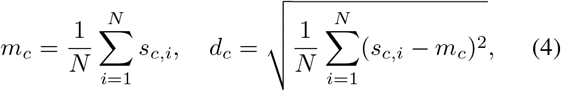

where *m*_*c*_ and *d*_*c*_ are the mean and standard deviation of channel *c*, respectively. This normalization ensured consistent feature scales, preserving the time column when present.

### B. Feature Extraction and Reduction

From each of the 20 selected EEG channels, 18 features were extracted, yielding 18× 20 = 360 features per subject. These features, listed in Table I, include statistical, Hjorth, band power, entropy, and wavelet metrics, capturing EEG signal characteristics for National Institutes of Health Stroke Scale (NIHSS) classification.

**Table I.** Extracted EEG Features.

| **Category** | **Features** |
|---|---|
| Statistical | Mean, Std, Variance, Skewness, Kurtosis, Median |
| Hjorth | Activity, Mobility, Complexity |
| Band Power | Delta, Theta, Alpha, Beta, Gamma |
| Entropy | Shannon Entropy |
| Wavelet | Wavelet Energy Percent (db4) |

Two techniques reduced the 360 features to 50:

#### SelectKBest Method

SelectKBest used ANOVA F-scores to select the top 50 discriminative features:

,
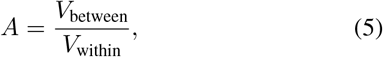

where *V*_between_ is variance between NIHSS classes, and *V*_within_ is variance within classes.

#### Principal Component Analysis (PCA)

PCA projected the standardized feature matrix onto 50 components:

,
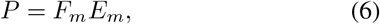

where *F*_*m*_ = standardized feature matrix, *E*_*m*_ = eigenvector matrix, and *P* is the projected matrix.

### C. Classification and Validation

Four classifiers—SVM, Random Forest, XGBoost, and KNN—were evaluated on both SelectKBest and PCA feature sets within a 5-fold cross-validation framework to ensure robust performance estimates. Stratified sampling preserved the NIHSS class distribution in each fold. The SVM with SelectKBest features demonstrated superior performance (as detailed in the results) and was selected for augmentation experiments.

### D. Data Augmentation Methods

To address class imbalance, particularly the underrepresentation of severe stroke cases, four augmentation techniques—SMOTE, ADASYN, WGAN, and WGAN-GP—were applied to the training folds during cross-validation. These methods generated synthetic samples for minority classes (moderate and severe), while test folds remained unaugmented to ensure unbiased evaluation.

### E. Generative Adversarial Networks

Generative Adversarial Networks (GANs) generated synthetic EEG features to balance minority classes (moderate and severe strokes). The GAN framework comprises a Generator (*G*_*n*_) and Discriminator (*D*_*s*_), trained adversarially to produce realistic samples from noise. WGAN and WGAN-GP were applied to training data in a 5-fold cross-validation to improve Support Vector Machine (SVM) performance for classes. Figure 2 shows the GAN architecture.

**Fig. 2.**
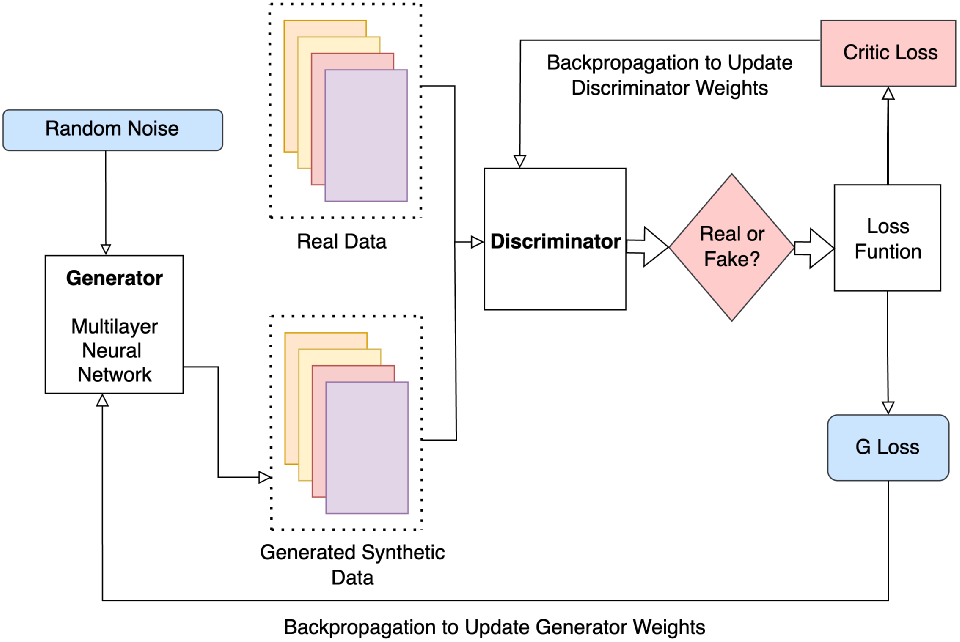
Architecture of a standard GAN.

#### SMOTE

Synthetic Minority Oversampling Technique (SMOTE) interpolates between minority class samples and their *k*-nearest neighbors to create synthetic samples. Applied to training data, SMOTE balanced classes, reducing bias toward mild strokes and boosting SVM accuracy for severe cases.

#### ADASYN

Adaptive Synthetic Sampling (ADASYN) prioritizes minority samples near the decision boundary. Used in each fold, ADASYN improved SVM sensitivity to severe stroke cases, enhancing performance for underrepresented classes.

#### WGAN

Wasserstein Generative Adversarial Network (WGAN) uses a Critic (*C*), where *C* = Critic, with Wasserstein distance for stable training via weight clipping. WGAN generated synthetic EEG features, improving SVM performance for minority classes.

#### WGAN-GP

WGAN with Gradient Penalty (WGAN-GP) uses a gradient penalty for enhanced sample quality. Applied in each fold, WGAN-GP produced realistic EEG features, significantly improving SVM performance across classes.

### F. Evaluation Metrics

Model performance was evaluated using macro-averaged metrics to address class imbalance in the categories (mild, moderate, severe). Macro-averaging ensures equal weighting of each class, enhancing fairness for underrepresented severe cases. Metrics were averaged across 5-fold cross-validation for robust estimates.

#### Accuracy

Measures the ratio of correct predictions across all classes:

,
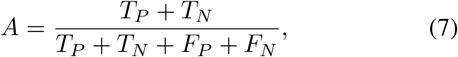

where *T*_*P*_ = true positives, *T*_*N*_ = true negatives, *F*_*P*_ = false positives, and *F*_*N*_ = false negatives.

#### Precision

Quantifies the proportion of correct positive predictions, macro-averaged for balanced class evaluation:

,
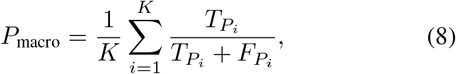

where *K* is the number of classes.

#### Recall

Assesses the proportion of actual positives identified, macro-averaged to prioritize minority classes:

,
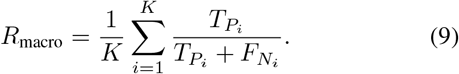

#### F1-Score

Computes the harmonic mean of precision and recall, macro-averaged for balanced performance:

,
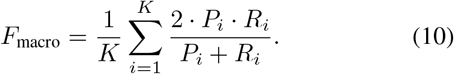

#### ROC-AUC

Evaluates the area under the receiver operating characteristic curve, measuring discriminative ability:

,
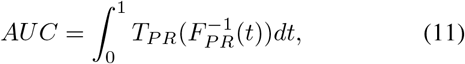

where *T*_*P R*_ = true positive rate and *F*_*P R*_ = false positive rate.

### G. Application to EEG Dataset

In each fold, training data was standardized, augmented using one technique (SMOTE, ADASYN, WGAN, or WGAN-GP), and used to train an SVM with an RBF kernel. A baseline SVM was trained on unaugmented data. Test sets remained unaugmented, and performance was evaluated using the above metrics, with confusion matrices and ROC curves generated per fold to assess classification robustness for class categories.

## IV. Results and Discussion

This section presents the experimental outcomes of the proposed EEG-based stroke severity classification framework. We first compare feature reduction techniques and classifiers, followed by an evaluation of data augmentation methods to address class imbalance.

### A. Feature Selection and Classifier Evaluation

Two feature reduction techniques—SelectKBest and Principal Component Analysis (PCA)—reduced the 360 EEG features to 50. Their performance was assessed using four classifiers: Support Vector Machine (SVM), Random Forest, XGBoost, and K-Nearest Neighbors (KNN) in a 5-fold cross-validation framework.

Table II shows the performance of classifiers on SelectKBest features. SVM achieved the highest accuracy (0.64), precision (0.434), recall (0.489), F1-score (0.429), and ROC-AUC (0.803), indicating superior discriminative ability for classes.

Figure 3 illustrates the 5-fold cross-validated accuracies for SelectKBest, with SVM showing the highest bar (0.64), confirming its effectiveness.

**TABLE II.**
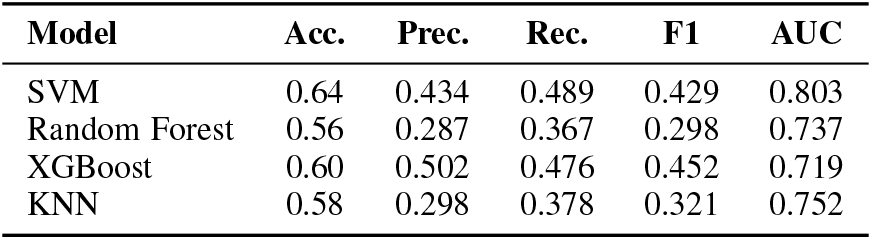
Model Performance on SelectKBest Features.

**Fig. 3.**
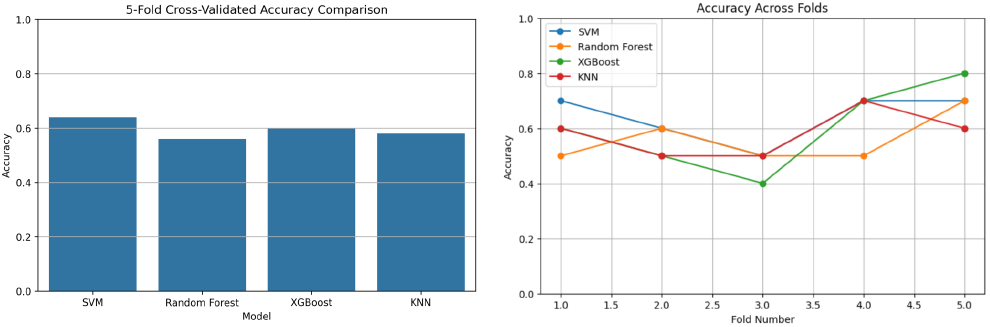
Accuracy Comparison on SelectKBest Features

Table III presents the performance on PCA features. Random Forest achieved the highest accuracy (0.56), but all models performed worse than with SelectKBest, with SVM at 0.54 and lower ROC-AUC (0.573), indicating reduced discriminative power.

**TABLE III.** Model Performance on PCA Features.

| **Model** | **Acc.** | **Prec.** | **Rec.** | **F1** | **AUC** |
|---|---|---|---|---|---|
| SVM | 0.54 | 0.184 | 0.322 | 0.233 | 0.573 |
| Random Forest | 0.56 | 0.278 | 0.367 | 0.300 | 0.631 |
| XGBoost | 0.44 | 0.276 | 0.327 | 0.296 | 0.616 |
| KNN | 0.50 | 0.182 | 0.298 | 0.225 | 0.594 |

Figure 4 shows the accuracies for PCA features, with Random Forest’s bar slightly higher (0.56) but overall lower than SelectKBest, highlighting PCA’s limitations.

**Fig. 4.**
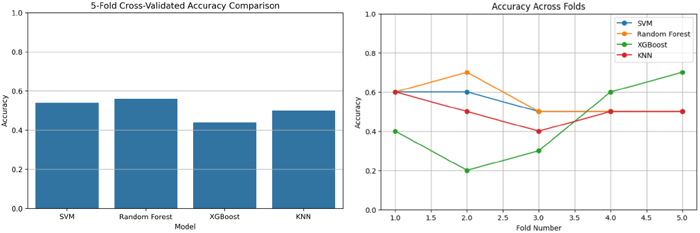
Accuracy Comparison on PCA Features

Table IV compares SelectKBest and PCA accuracies, showing SelectKBest’s superiority in three of four models, with SVM on SelectKBest achieving the highest accuracy (0.64). Thus, SelectKBest with SVM was selected for augmentation experiments.

**TABLE IV.** Accuracy Comparison: SelectKBEST VS PCA.

| **Model** | **SelectKBest Acc.** | **PCA Acc.** |
|---|---|---|
| SVM | 0.64 | 0.54 |
| Random Forest | 0.56 | 0.56 |
| XGBoost | 0.60 | 0.44 |
| KNN | 0.58 | 0.50 |

### B. Augmentation Technique Results

To address class imbalance in severity labels (mild, moderate, severe), four augmentation techniques—SMOTE, ADASYN, WGAN, WGAN-GP—were applied to training folds in a 5-fold cross-validation setup using the selected SelectKBest feature selection followed by SVM configuration.

Test folds remained unaugmented to ensure unbiased evaluation, focusing on improving classification performance for minority classes.

Table V presents SVM performance with and without SMOTE. SMOTE significantly enhanced all metrics, achieving a precision of 0.72 and recall of 0.67, reflecting its ability to generate synthetic samples that improve detection of moderate and severe classes.

**TABLE V.** Performance: With vs Without SMOTE.

| Metric | Without SMOTE | With SMOTE |
|---|---|---|
| Acc. | 0.64 | 0.70 |
| Prec. | 0.42 | 0.72 |
| Rec. | 0.47 | 0.67 |
| F1 | 0.42 | 0.67 |
| AUC | 0.84 | 0.87 |

Table VI shows ADASYN’s performance, with moderate improvements over the baseline, particularly in precision (0.58) and recall (0.55), indicating effective augmentation for challenging minority class samples near decision boundaries.

**TABLE VI.** Performance: With VS Without ADASYN.

| Metric | Without ADASYN | With ADASYN |
|---|---|---|
| Acc. | 0.64 | 0.66 |
| Prec. | 0.42 | 0.58 |
| Rec. | 0.47 | 0.55 |
| F1 | 0.42 | 0.54 |
| AUC | 0.84 | 0.86 |

Table VII displays WGAN’s results, with improved accuracy (0.68) but a lower AUC (0.82) compared to SMOTE and ADASYN, suggesting moderate success in generating realistic synthetic EEG features for minority classes.

**TABLE VII.**
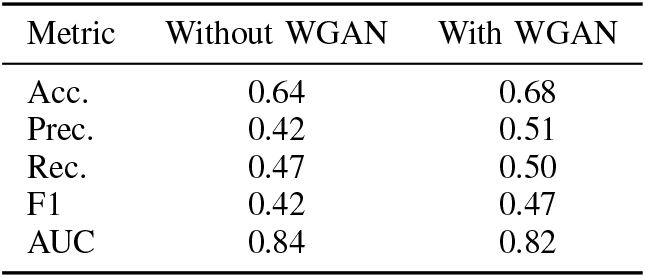
Performance: With vs Without WGAN.

| Metric | Without WGAN | With WGAN |
|---|---|---|
| Acc. | 0.64 | 0.68 |
| Prec. | 0.42 | 0.51 |
| Rec. | 0.47 | 0.50 |
| F1 | 0.42 | 0.47 |
| AUC | 0.84 | 0.82 |

Table VIII illustrates WGAN-GP’s performance, achieving the highest accuracy (0.72) and balanced metrics, including recall (0.58) and AUC (0.85), highlighting its ability to produce high-quality synthetic samples for improved classification.

**TABLE VIII.**
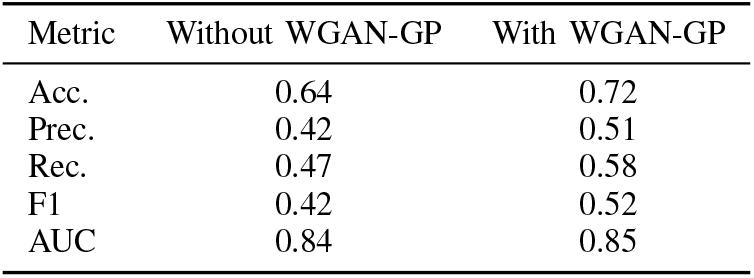
Performance: With vs Without WGAN-GP.

| Metric | Without WGAN-GP | With WGAN-GP |
|---|---|---|
| Acc. | 0.64 | 0.72 |
| Prec. | 0.42 | 0.51 |
| Rec. | 0.47 | 0.58 |
| F1 | 0.42 | 0.52 |
| AUC | 0.84 | 0.85 |

### C. Performance Comparison

Table IX summarizes the performance across augmentation methods. SMOTE exhibited strong performance in precision (0.72), recall (0.67), and F1-score (0.67), reflecting its robustness in addressing class imbalance. WGAN-GP achieved the highest accuracy (0.72) and balanced overall metrics (recall 0.58, AUC 0.85), making it the most effective for classification due to its stable generation of synthetic EEG features.

**TABLE IX.** Comparison of Augmentation Methods.

| Method | Acc. | Prec. | Rec. | F1 | AUC |
|---|---|---|---|---|---|
| Without Aug. | 0.64 | 0.42 | 0.47 | 0.42 | 0.84 |
| SMOTE | 0.70 | 0.72 | 0.67 | 0.67 | 0.87 |
| ADASYN | 0.66 | 0.58 | 0.55 | 0.54 | 0.86 |
| WGAN | 0.68 | 0.51 | 0.50 | 0.47 | 0.82 |
| WGAN-GP | 0.72 | 0.51 | 0.58 | 0.52 | 0.85 |

Figure 5 illustrates ROC-AUC curves for SMOTE, ADASYN, WGAN, and WGAN-GP, with SMOTE’s curve reaching the highest AUC (0.87), followed closely by WGAN-GP (0.85), demonstrating their strong discriminative ability for NIHSS classes.

**Fig. 5.**
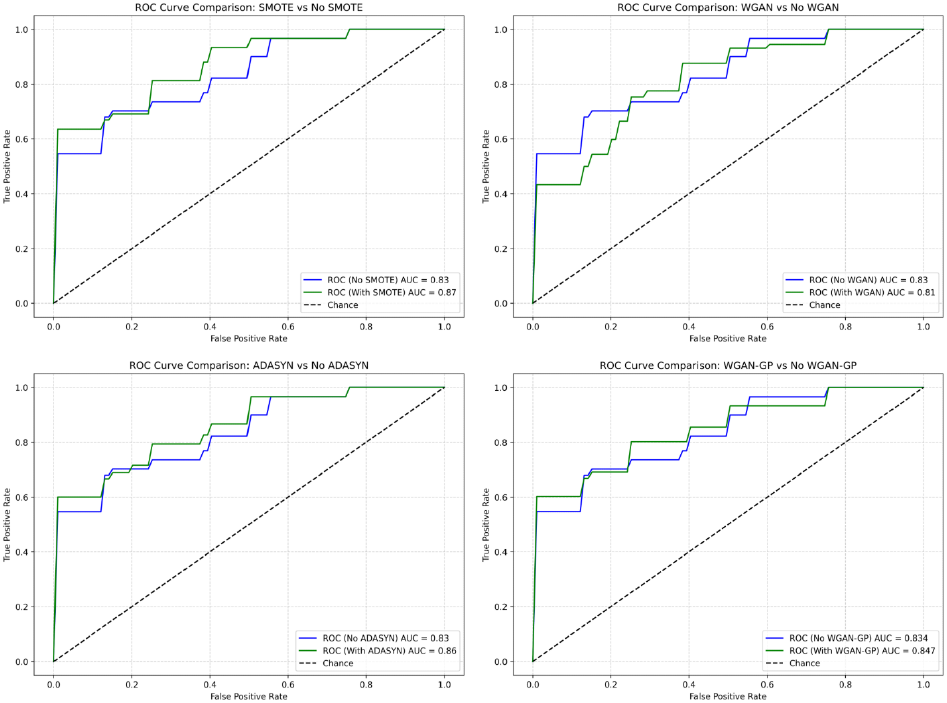
ROC-AUC Curves for Augmentation Methods

Figure 6 compares accuracies across methods, with WGAN-GP’s bar peaking at 0.72, followed by SMOTE at 0.70, visually confirming WGAN-GP’s superior accuracy for EEG-based stroke severity classification.

**Fig. 6.**
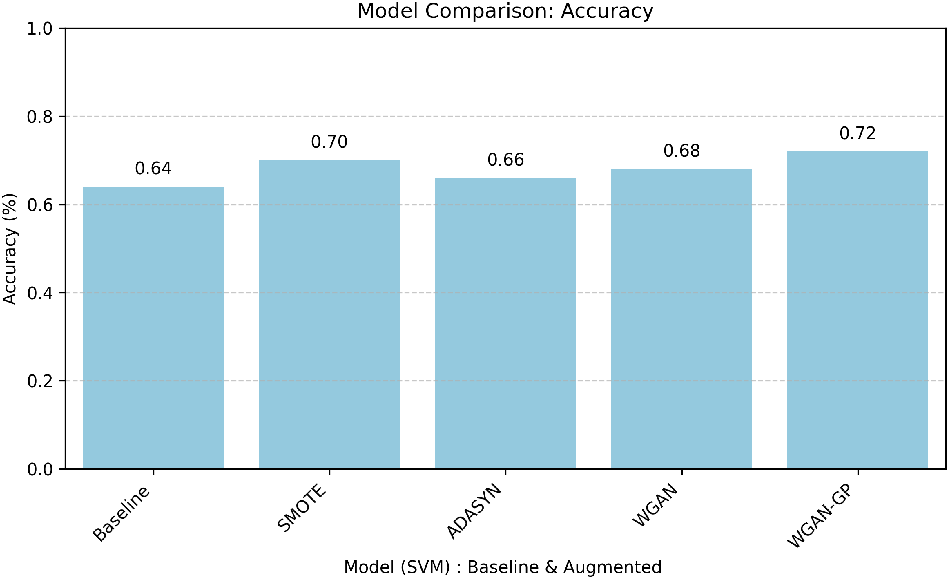
Accuracy: Baseline vs Augmentation Methods

### D. Key Insights

Our experiments highlight that combining SelectKBest feature selection with SVM yields strong baseline performance. Incorporating data augmentation further boosts accuracy, with WGAN-GP showing the best overall improvement. This suggests that a hybrid approach of feature selection, SVM classification, and generative augmentation offers a practical solution for improving EEG-based stroke severity classification.

## V. Conclusion

This study tackled stroke severity classification using EEG data, addressing challenges of limited samples and class imbalance, with feature selection using SelectKBest followed by evaluation of multiple classifiers identifying SVM as the most effective, and four data augmentation techniques—SMOTE, ADASYN, WGAN, WGAN-GP—applied to enhance model robustness, where WGAN-GP followed by SVM followed by SelectKBest achieved the highest accuracy (0.72), marking this approach among the first to apply generative augmentation for EEG-based stroke classification, demonstrating WGAN-GP’s superior performance over traditional methods, with the pipeline of SelectKBest feature selection followed by SVM followed by WGAN-GP providing a robust baseline for future EEG-based diagnostic research, while future work will validate these results on larger, more diverse datasets, explore deep learning models for raw EEG data, investigate hybrid augmentation techniques, and incorporate temporal modeling with recurrent neural networks or Transformers to further improve stroke severity assessment and clinical decision-making.

## Data Availability

The dataset supporting the conclusions of this article is publicly available in the Figshare repository: https://doi.org/10.6084/m9.figshare.21679035.v5

https://doi.org/10.6084/m9.figshare.21679035.v5

## Notes

### Competing Interest Statement

The authors have declared no competing interest.

### Author Declarations

The study used openly available human EEG data that were publicly accessible before the initiation of the study. The dataset was originally made available by Liu et al. on Figshare and can be accessed at: https://doi.org/10.6084/m9.figshare.21679035.v5

